# Early palliative care for patients with oral cancer in Sri Lanka: A quasi-experimental study

**DOI:** 10.1101/2025.03.05.25323404

**Authors:** Nadisha Ratnasekera, Irosha Perera, Sumeth Perera, Jenny Lau, Camilla Zimmermann, Pushpakumara Kandapola Arachchige

**Author notes:** **Corresponding Author information** Full Name - Nadisha Ratnasekera, E-mail address –.

## Abstract

**Objectives:** Oral cancer is prevalent among Sri Lankan men and compromises their quality of life (QOL). Limited research exists on providing early palliative care in low- to middle-income countries. We assessed the effectiveness of an early palliative care intervention in improving psychological distress and QOL among patients with oral cancer in Sri Lanka.

**Design:** A quasi-experimental study

**Setting:** The study took place at three tertiary care units providing oral cancer treatment in Sri Lanka: the oral and maxillofacial wards at the National Dental Hospital, Colombo (Teaching); the oral and maxillofacial wards at Colombo South (Teaching) Hospital, Karapitiya; and the onco-surgery wards at the National Cancer Institute Maharagama (Apeksha Hospital).

**Participants:** The study participants were patients with oral cancer whose definitive diagnosis had been communicated to the patient and had psychological distress (score of ≥4 after screening with the Sinhala version of the Distress Thermometer); awaiting surgery as the first treatment modality; and married with children, including at least one family caregiver with the ability to communicate and read well in Sinhalese. Exclusion criteria were, recurrent oral cancer; a formal psychiatric diagnosis; and receiving or having received any early palliative care intervention. They were divided to 55 controls and 55 cases based on the availability of an accessible Public Health Nursing Officer.

**Interventions:** The early palliative care package, comprising six components, was delivered in three sessions by the principal investigator (PI) and Public Health Nursing Officers.

**Outcome measures:** The effectiveness was assessed by the level of psychological distress using the Sinhala version of the Distress Thermometer and Level of quality of life using EORTC QLQ 30 with module H&N 35 at baseline (T0), post-intervention (T1), one month (T2), and three months (T3).

**Results:** In the intervention group, Distress Thermometer scores significantly decreased from baseline (T0) to three months post-intervention (T3) (6.9 vs. 3.5, p=0.0001). The QOL dimensions improved at all time points for both groups, with the intervention group showing a significantly higher improvement (p=0.0001).

**Conclusion:** The novel early palliative care intervention improved the psychological distress and quality of life of patients with oral cancer.

## Introduction

Oral cancer is one of the most common cancers in the world^1,2,3^, and the most common cancer in Sri Lankan men^4^, with a profound effect on the quality of life of patients and their families. After a diagnosis of oral cancer, patients experience not only disturbance of physical appearance and basic human functions such as eating, speech, and breathing^5,6,7^, but also emotional, social, and spiritual problems^8,9^. Psychological distress can adversely affect compliance with medical care, recovery from illness, adjustment to life after treatment, and survival^10-14^.

The World Health Organization defines palliative care as, “an approach that improves the quality of life of patients and their families who are facing problems associated with life-threatening illness. It prevents and relieves suffering through the early identification, correct assessment, and treatment of pain and other problems”^15^. In high-income countries, randomized controlled trials have shown that early palliative care for patients with advanced cancer improves the quality of life, mood, satisfaction with care, and symptom control^16-19^; this evidence has led to recommendations by major cancer organizations that early palliative care be enacted for all patients with advanced cancer^20^. In addition, the National Comprehensive Cancer Network (NCCN) recommends routine screening for distress among all patients with cancer and considers psychological distress as the sixth vital sign of cancer care^14^. However, neither distress screening nor integration of palliative care are routinely available in low- and middle-income countries (LMICs), and there has been scant prospective research on the impact of early palliative care in these countries^21^.

In the Sri Lankan health system, inpatient cancer care is provided mainly through the public sector, which provides universal access to cancer treatment, and outpatient cancer care is delivered through both the public and private sectors^22^. Sri Lanka has a national strategic framework for palliative care development (from 2018 to 2023), which includes pain and symptom management; psychological, emotional, social, and spiritual support; and support of family and caregiver coping during the patient’s illness and bereavement period^23^. Palliative care is mainly delivered by oncologists and consultant psychiatrists in the 25 cancer centres across Sri Lanka^24^. The only specialized palliative care clinic is at the National Cancer Institute Sri Lanka - Apeksha Hospital, headed by a consultant oncologist^24^. In 2018, a postgraduate diploma program in Palliative Medicine was introduced to address the need for professionals in palliative care in Sri Lanka^24^. Despite these advances, palliative care for patients with cancer is provided only at the end of life and there is no special team assigned to provide palliative care in Sri Lanka^24,25^.

The aim of this study was to determine the effectiveness of an early palliative care intervention in improving psychological distress and quality of life among patients with oral cancer in Sri Lanka.

## Methods

This study was reported in accordance with the TREND statement (Appendix I)^26^.

### Study Design and Participants

This was a multi-center quasi-experimental study, where an intervention group and a control group were present but randomization was not carried out. The study took place at three tertiary care units providing oral cancer treatment in Sri Lanka: the oral and maxillofacial wards at the National Dental Hospital, Colombo (Teaching); the oral and maxillofacial wards at Colombo South (Teaching) Hospital, Karapitiya; and the onco-surgery wards at the National Cancer Institute Maharagama (Apeksha Hospital). Inclusion criteria were: definitive diagnosis communicated to the patient; presence of psychological distress (score of ≥4 after screening with the Sinhala version of the Distress Thermometer^27^); awaiting surgery as the first treatment modality; and married with children, including at least one family caregiver with the ability to communicate and read well in Sinhalese. Exclusion criteria were: recurrent oral cancer; a formal psychiatric diagnosis; and receiving or having received any early palliative care intervention. All study procedures were in accordance with the ethical standards of the Ethics Board, Faculty of Medicine, University of Colombo, Sri Lanka (registration number EC-18-097) and with the 1964 Helsinki Declaration and its later amendments or comparable ethical standards.

### Study Procedures

All patients who fulfilled the eligibility criteria were approached for recruitment. The PI met patients at the ward when they were admitted for the surgery. The eligible patients were introduced to the study and the information sheet, and the consent form was given before study commencement. Informed consent was obtained from all individual participants included in the study. Each participant was given an index number and a patient’s record form.

The participants were divided into two groups, based on the availability of an ‘accessible Public Health Nursing Officer’. The Public Health Nursing Officer is a new healthcare category in Sri Lanka, assigned to Healthy Lifestyle Centres in some of the island’s base hospitals^28^. Public Health Nursing Officers are expected to carry out both field and hospital services. There are four key areas in their job description: non-communicable disease activities, palliative care activities, geriatric care, and provision of specific services such as disaster management. Palliative care activities are focused on providing relief from symptoms and stress of chronic illness, with the goal of improving quality of life for both the client and the family^28^. The ‘accessible Public Health Nursing Officer’ was defined as, “a Public Health Nursing Officer who could reach the residence of the eligible patient within 1.5 hours (maximum). The mode of transportation could be by personal/public transport”. Eligible patients with oral cancer who lived in an area where an accessible Public Health Nursing Officer was present were assigned to the intervention group and received the early palliative care intervention package. Eligible patients with oral cancer who lived where an accessible Public Health Nursing Officer was not present were assigned to the control group.

### Intervention Group

The intervention was delivered by the PI and the Public Health Nursing Officers, whose capacity to deliver the intervention package was enhanced through a training programme^29,30^. The training programme was developed with the guidance of experts in the fields of oncology, Oro-maxillo-facial surgery, psychiatry, sociology, and psychology which consisted of two sessions. The first session was a 2-day in-person training session given to all Public Health Nursing Officers in Sri Lanka. The second session was online and was specifically designed for Public Health Nursing Officers participating in session 1 who were selected to deliver the intervention package. The evaluation of the training programme was carried out according to the Kirkpatrick Training Evaluation Model^31^. A handbook was provided for the Public Health Nursing Officers with instructions on how to carry out the intervention.

The early palliative care intervention package was developed following the guidelines provided by the UK Medical Research Council for the development of complex interventions^32^. The early palliative care intervention for patients with oral cancer had six components: 1) providing information, 2) addressing acute and functional issues, 3) nutritional care, 4) psychological support, 5) mindfulness therapy, and 6) coordination of the financial allowance. Appendix II gives comprehensive details of the intervention and Appendix III provides the TIDieR checklist^33^. The package was formulated by triangulating literature review findings and several independent studies: a case-control study; in-depth interviews with experts in the field of oncology and palliative care in Sri Lanka; key-informant interviews with patients with oral cancer and their caregivers; and an observation study at the National Cancer Institute Sri Lanka-Apeksha Hospital)^29^. The intervention package was finalized through a nominal group discussion with experts in the fields of oncology, oro-maxillo-facial surgery, psychiatry, sociology, and psychology.

The intervention was delivered by the PI and Public Health Nursing Officer in 3 sessions. The first session of the intervention was delivered by the PI, immediately after the patient provided consent. It was delivered at the ward after the patient was admitted for surgery. After delivering the first session, the PI contacted the relevant Public Health Nursing Officer in the area of the patient’s residence and handed over the patient to him/her to carry out the second and third sessions at the patient’s residence, 1 week and 3 weeks, respectively, after the patient was discharged from the hospital. Each session averagely took 2 hours.

Certain study parameters were undertaken to ensure proper delivery of the early palliative care intervention package. To maintain the uniformity of the intervention, a handbook was provided to the Public Health Nursing Officers in the area of each patient’s residence, with specific guidance to deliver the intervention^29^. In addition, there was close monitoring of the Public Health Nursing Officers on delivering the intervention package by contacting them thrice during the delivery of the intervention. To improve the compliance of the patients, the follow-up appointments were scheduled according to patients’ convenience and only on their clinic days. Patients were reminded twice about appointments before the date.

### Control group

The control group received standard care currently existing in the health system where the patient has access to some of the palliative care services upon the patient’s request. However, these services are provided in an ad hoc manner and mainly focused on relieving physical symptoms, and no special focus on palliative care. Table 1 shows the comparison of the early palliative care intervention and the standard care received by the intervention and control groups, respectively.

**Table 1:** Comparison of the early palliative care intervention package and standard care.

| Description | Early palliative care | Standard Care |
| --- | --- | --- |
| Care in the ward<br>(Once the patient got admitted to the ward for the surgery) | Routine oncology care and the PI visited to deliver the first session of the intervention package | Routine oncology care only |
| At the discharge | Routine oncology care<br>Patient was provided with an information-providing booklet and a dietary plan after the consultation with the dietician in the hospital.<br>The patient was connected to the Public Health Nursing Officer of the area | Routine oncology care only. Based on the oncologist's decision some patients receive a consultation with the dietician. |
| Home care | Public Health Nursing Officer visited the patient's residence twice with a two-week gap and delivered the last two sessions of the intervention | No home care |
| Components of the intervention package |  |  |
| Providing information | Routine method<br>An information-providing booklet and a video clip were given<br>In-person discussion with the PI and Public Health Nursing Officer | Routine method - talking to the medical/dental officers and nursing officers, during clinic dates and ward rounds |
| Addressing acute and functional issues | Routine method<br>Direct access to the PI and the Public Health Nursing Officer<br>Hassle free referrals for pain and other acute issues management | Routine method - seeking help from the medical/dental officers and nursing officers, during clinic dates and ward rounds |
| Nutritional care | Routine method<br>A consultation with the dietician (after the surgery before the discharge from the ward).<br>Provision of a customized diet plan<br>Follow-up on the compliance to the diet plan and support through the Public Health Nursing Officer | Routine method – ad-hoc. Referral to the dietician depends on the oncologist’s decision. If the patient complained of some difficulty in taking food it was addressed. |
| Psychological support | Problem-solving counseling and follow up care | No special support |
| Mindfulness therapy | Mindfulness based therapy and follow up care by the PI and the Public Health Nursing Officer. | No therapy |
| Coordination of the financial allowance | The PI and the Public Health Nursing Officer reached out to the social worker at the relevant District Secretariat office and made sure the financial allowance was received without delay or any hassle. | Only patient’s attempts to connect with the social worker at the District Secretariat |

### Outcome measures

Data were collected by the PI and the Public Health Nursing Officers. The outcome measures were collected at four-time points: at enrollment (T0) (by the PI at the tertiary care hospital); immediately after delivering the intervention package (T1) (by the Public Health Nursing Officer at the patient’s residence); and 1 month (T2) and 3 months (T3) after the intervention (by the PI at the tertiary care hospital during clinic visits).

The outcomes were the level of psychological distress and quality of life of the patient. The level of psychological distress was measured through the mean scores of the Sinhala version of the Distress Thermometer^27^. The Distress Thermometer is 1 item, an ultra-short, visual analogue screening tool, originally developed in the USA for prostate cancer patients in 1998^34^. The Distress Thermometer comes with a Problem List. This has 35 items and it covers 5 life domains (practical, family/social, emotional, spiritual, and physical problems)^35^. The Distress Thermometer and Problem List were translated into the Sinhala language (the primary language in Sri Lanka) and cross-culturally adapted to Sri Lanka^27^.

Quality of life was measured using the Sinhala version of the EORTC QLQ 30 and H&N 35^36^. The original measures were developed by The European Organization for Research and Treatment of Cancer (EORTC) Quality of Life Group (QLG)^37^. These tools were translated and validated to Sri Lanka through a large-scale multicentre study^36^.

Additionally, process evaluation indicators measured the reception of the intervention package within and between the groups; these are presented in Appendix IV.

### Statistical Methods

The sample size was calculated using a minimally important difference of 2 of quality of life and a standard deviation of 4^38^, assuming a power of 80% and a type I error of 0.1 two-sided. After accounting for non-response rate of 5-10%, the target sample size was 55 per group or 110 in total.

All analyses were performed by intention to treat, with analysis within the groups to which patients had originally been assigned, regardless of whether or not they actually received their intended intervention. SPSS version 21 software package was used for data analysis. The baseline sociodemographic data, socioeconomic data, and other selected characteristics were compared between the intervention group and the control group.

Distress Thermometer scores and quality of life subscale scores were compared within and between the intervention group and the control groups. The EORTC QLQ 30-H&N 35 subscale scores were calculated as per the EORTC QLQ-Q30 Scoring Manual^37^. The scores distributions were assessed for normality. Since the data was in a skewed distribution, the Mann-Whitney U test and Friedmann ANOVA were used to compare the change of mean scores. As multiple comparisons were present, the Bonferroni correction was carried out. When statistical significance was detected across multiple time points, the Wilcoxon Signed Rank Test was applied to pinpoint the specific time points of significance. Linear regression was carried out to control for the potential confounders and assess the impact of the novel intervention on psychological distress and quality of life.

## Results

The study took place from October 2019 to May 2020. Of 223 patients who were screened, 110 were eligible and provided follow-up data for all time points, with a response rate of 100%. Although there was no loss to follow-up, 12 participants of the intervention group and 5 in the control group had delayed delivery of the intervention and delayed data collection due to the COVID-19 pandemic (Figure 1).

**Figure 1:**
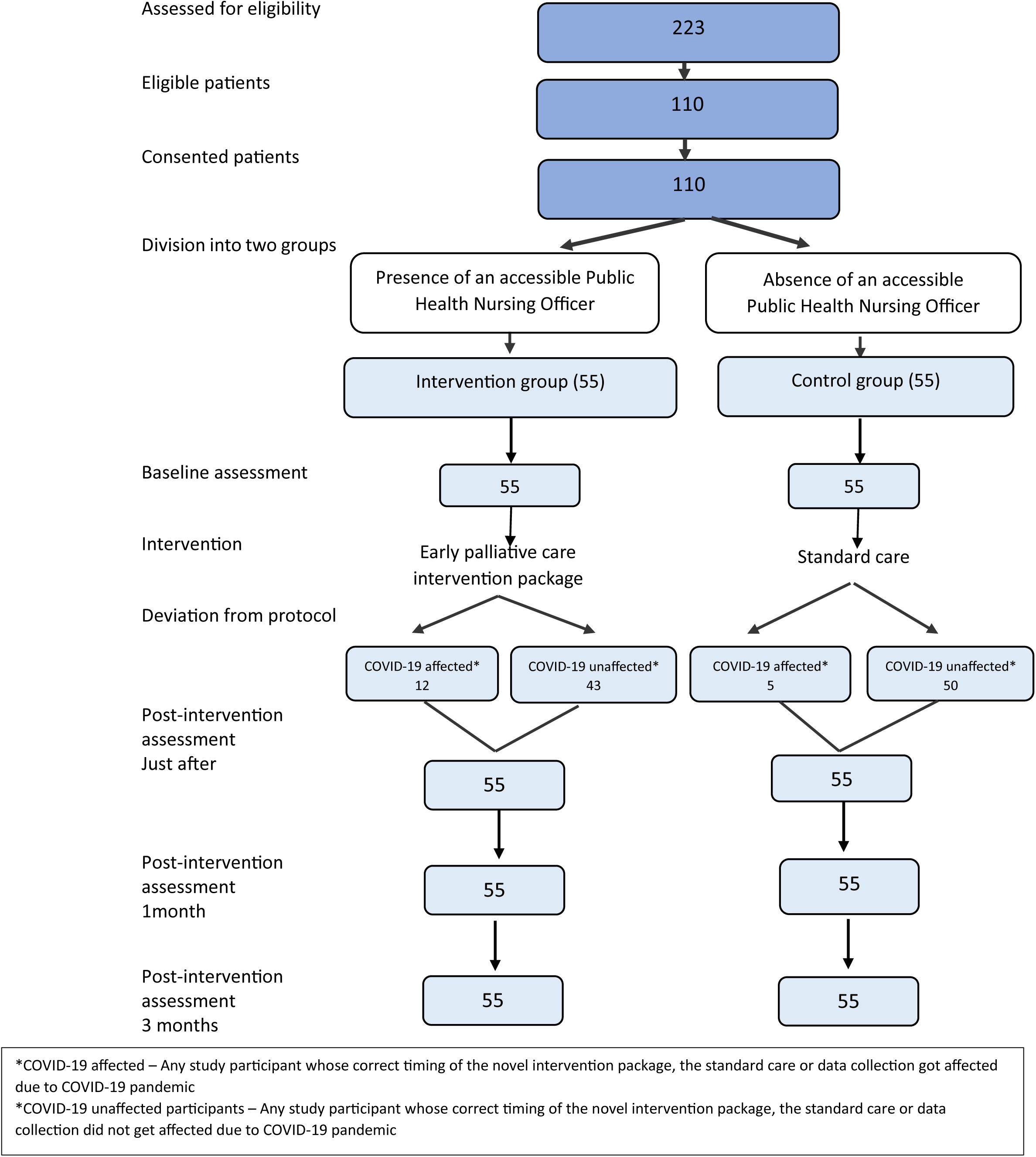
Patients progress through the study – flow chart

As per the planned intention-to-treat analysis, all participants (whether or not the intervention and data collection were affected due to COVID-19) were analyzed within the study group that they were originally assigned. Asub-group analysis revealed no significant difference between the outcomes of the COVID -19 affected participants and unaffected participants (Appendix V).

### Sample characteristics

Table 2 shows the socio-demographic, socio-economic and medical characteristics of the two groups, which were similar at baseline.

**Table 2:** Comparison of the selected characteristics of the intervention group and control group.

| Characteristic | IG<sup>a</sup> (n= 55)<br>No. (%) | CG<sup>b</sup> (n= 55)<br>No. (%) | Total (n=110)<br>No. (%) | P value* |
| --- | --- | --- | --- | --- |
| Selected socio-demographic characteristics |  |  |  |  |
| Age (Years)- Mean = 56.72 (SD- 10.57), Median = 58.00 (Range= 38 - 76) |  |  |  |  |
| 30-40 | 7 (12.7) | 4 (7.3) | 11 (10) | 0.41 |
| 41-59 | 27 (49.1) | 24 (43.6) | 51 (46.4) |  |
| 60-80 | 21 (38.2) | 27 (49.1) | 48 (43.6) |  |
| Sex |  |  |  |  |
| Male | 43 (78.2) | 42 (76.4) | 85 (77.3) | 0.82 |
| Female | 12 (21.8) | 13 (23.6) | 25 (22.7) |  |
| Ethnicity |  |  |  |  |
| Sinhala | 47 (85.4) | 43 (78.2) | 90 (81.8) | 0.47 |
| Tamils | 4 (7.3) | 8 (14.5) | 12 (10.9) |  |
| Moors and others | 4 (7.3) | 4 (7.3) | 8 (7.3) |  |
| Selected socio-economic characteristics |  |  |  |  |
| Level of education |  |  |  |  |
| No schooling | 4 (7.3) | 9 (16.4) | 13 (11.8) | 0.34 |
| Primary (up to grade 5) | 18 (32.7) | 16 (29.1) | 34 (3.1) |  |
| Secondary (10 and up) | 33 (60.0) | 30 (54.5) | 63 (57.3) |  |
| Occupation category |  |  |  |  |
| Unemployed | 23 (41.8) | 33 (60.0) | 56 (50.9) | 0.073 |
| Skilled<sup>a</sup> and unskilled<sup>b</sup> | 27 (49.1) | 21 (38.2) | 48 (87.3) |  |
| Professional and clerical | 5 (9.1) | 1 (1.8) | 6 (10.9) |  |
| Selected medical characteristics |  |  |  |  |
| Treated tertiary care institute |  |  |  |  |
| Apeksha Hospital | 15 (27.3) | 10 (18.2) | 25 (23.7) | 0.84 |
| National Dental Hospital | 13 (23.6) | 23 (39.9) | 36 (32.7) |  |
| Karapitiya Teaching Hospital | 17 (30.9) | 17 (30.9) | 34 (30.9) |  |
| Colombo South Teaching Hospital | 5 (9.1) | 10 (14.9) | 15 (13.6) |  |

| **Stage of the disease** |  |  |  |  |
| --- | --- | --- | --- | --- |
| Early | 17 (30.9) | 16 (29.1) | 33 (30.0) | * 0.24 |
| Late | 38 (69.1) | 39 (70.1) | 77 (70.0) |  |

(a= Intervention group, b= Control group, \*=Chi Square, c=Presence of the local lesion only, d= the lymph nodes are involved, a- labourers who have undergone a short training, b- labourers who have not undergone any training)

### Level of psychological distress

The level of psychological distress was measured through the Distress Thermometer scores where the data set showed a skewed distribution. The comparison of the Distress Thermometer scores within and between the intervention group and control group at T0, T1, T2, and T3 is presented in Figure 2.

**Figure 2:**
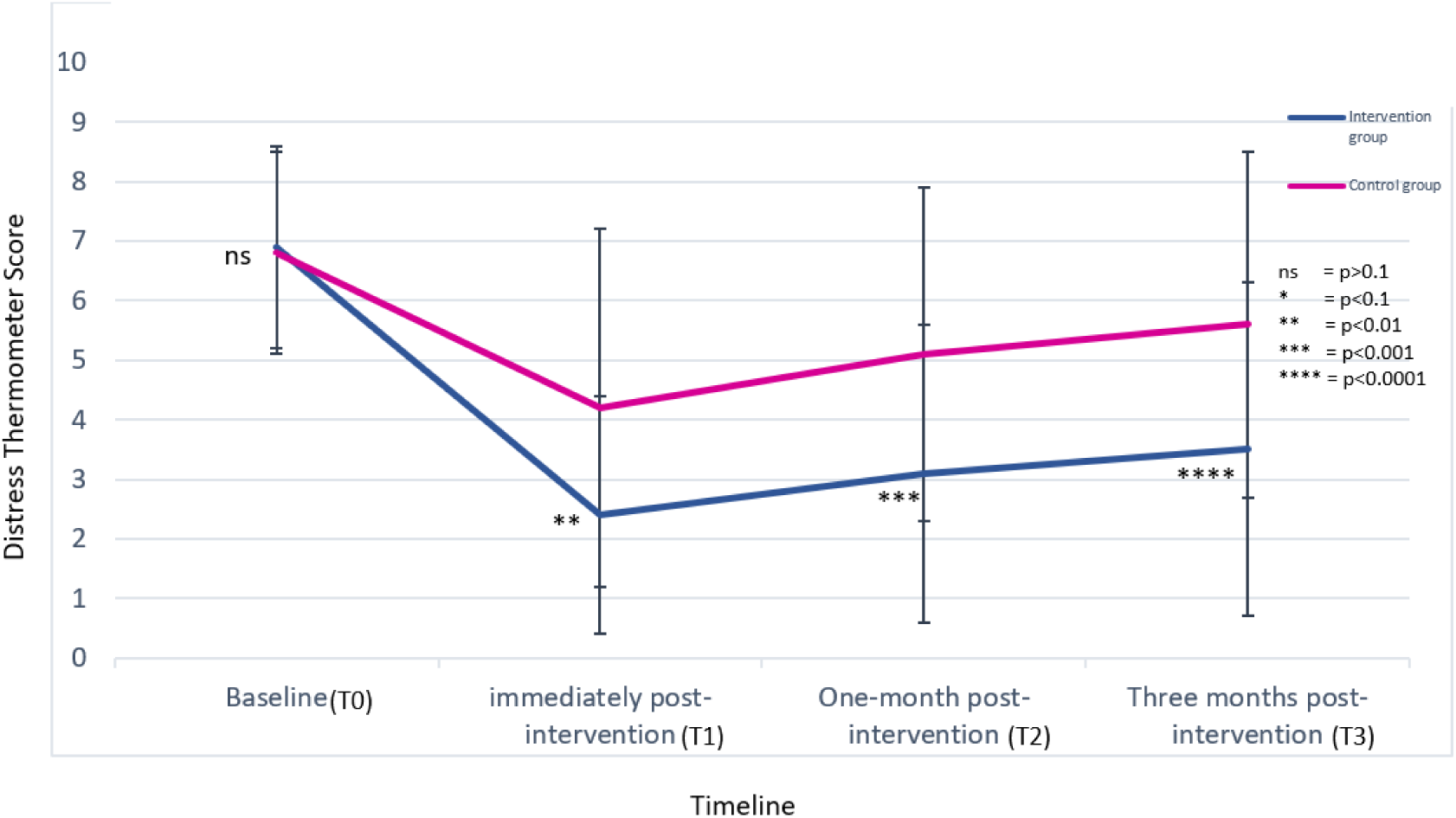
Comparison of the Distress Thermometer mean scores, between and within intervention and control groups

According to Figure 2, the Distress Thermometer mean scores showed no significant difference at the baseline between the intervention group and control group. In all post-intervention assessments, the Distress Thermometer mean score was lower in the intervention group than the control group and was statistically significant (T1 = 2.4 [2.0] vs. 4.2 [3.0] p= 0.001, T2 = 3.1[2.5] vs. 5.1[2.8] p=0.001, T3 = 3.5[2.8] vs. 5.6[2.9] p=0.0001).

### Quality of Life

We compared the quality of life sub-scale scores (Global health status-GHS, Functional Scales-FS, Symptoms Scales-SS and H&N 35 module) in both the intervention group and control group pre- and post-intervention (Figure 3).

**Figure 3:**
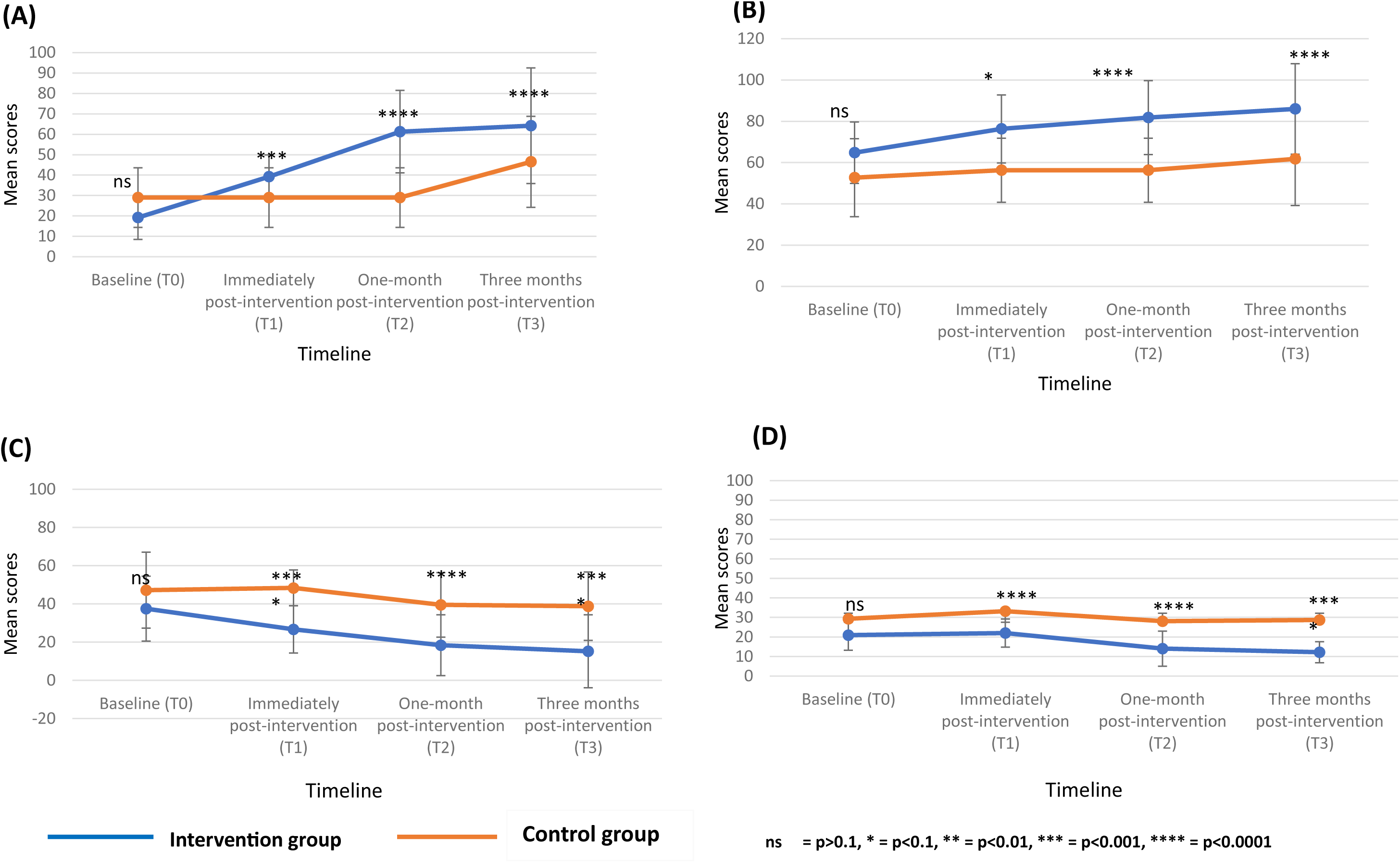
Comparison of the scores of (A) Global Health Status/ Quality of Life, (B) between Functional Status (C) Symptom Status, (D) H&N 35 Module between the intervention group and control group,

There was no significant difference in the baseline mean scores of all quality of life dimensions between the intervention group and control groups. There was a higher mean score of all dimensions of quality of life in the intervention group compared to the control group at all post-intervention time points (GHST1= 39.2[10.7] vs. 29[14.6], GHST2 = 61.3[20.2] vs. 29[14.6], GHST3 = 64.2 [28.3] vs. 46.5[22.3], FST1 = 76.3[16.5] vs. 56.3[15.5], FST2 = 81.8[17.9] vs. 56.2[15.5], FST3 = 86[21.9] vs. 61.8[22.6], SST1 = 26.7[12.4] vs. 48.4[9.4], SST2 = 18.4[15.9] vs. 39.5[16.9], SST3 = 15.2[19.1] vs. 38.8[17.9]).

### Multivariable Analysis

After controlling for confounding factors, the Distress Thermometer score was 2.26 (0.57) points lower in the intervention group compared to the control group at T3 (Table 3).

**Table 3:** Effectiveness of the intervention on level of psychological distress after controlling for confounders.

| Factor | $\beta$(SE) | 95% CI for $\beta$ | | P value |
| --- | --- | --- | --- | --- |
|  |  | Lower | Upper |  |
| Level of psychological distress at the baseline (T0) | 0.22 (0.16) | -0.12 | 0.53 | 0.17 |
| Age | 0.01 (0.04) | -0.08 | 0.05 | 0.2 |
| Female | 0.20 (0.69) | -1.16 | 1.58 | 0.62 |
| Sinhala | 0.16 (0.51) | -0.85 | 1.17 | 0.89 |
| Secondary education | 0.10 (0.59) | -0.79 | 0.57 | 0.75 |
| Unemployment | 0.27 (0.72) | -0.89 | 1.44 | 0.50 |
| Late stage | 1.30(0.74) | -0.13 | 2.75 | 0.05 |
| **Intervention group** | **-2.26 (0.57)** | **-1.07** | **-3.35** | **0.0001** |

After adjusting for confounding variables, the sub-dimensions of quality of life demonstrated significant differences at T3 in the intervention group compared to the control group: Functional Status was 23.6 (4.5) higher, Global Health Status 17.1 (5.4) points higher, Symptom Status 23.2 (3.6) lower.

## Discussion

This study provides evidence to support that the early palliative care intervention package significantly improves psychological distress and the quality of life of patients with oral cancer just after, 1 month after, and 3 months after the intervention compared with standard cancer care.

To the best of our knowledge, this is the first study on providing early palliative care for patients with oral cancer in Sri Lanka. The systematic review carried out in 2008 discovered 22 trials on specialized palliative care from 1984 to 2007 half of which were exclusively on cancer patients^39^. It suggested that evidence supporting the effectiveness of specialized palliative care was sparse and limited by methodological shortcomings. In our study, we attempted to address these issues by reducing control group contamination and attrition. However, we selected a quasi-experimental study design since this is the first study of this nature in Sri Lanka and a randomized control trial could be planned based on the findings of this study.

The formulation of the novel intervention package followed a thorough scientific process stipulated by the UK Medical Research Council for the development of complex interventions which assured the scientific rigor of the intervention package^32^. The intervention was a multi-component package that addressed six of the eight domains identified by the National Consensus Project on Clinical Practice Guidelines for Quality Palliative Care^40^. Especially, the intervention package utilized the existing resources in the health system by including Public Health Nursing Officers (PHNOs) who were already involved in the healthcare delivery. This favors the implementation and sustainability of the novel package in the low-resource health system in Sri Lanka.

Our selection of quality of life as one of the outcomes of this study is supported since the ultimate goal of providing palliative care is to improve quality of life^15^. Selecting the psychological distress of the patients as the other main outcome of interest could be justified as there are proven adverse effects of untreated psychological distress on the cancer journey^5-7^.

The key findings of our study show similar improvements in terms of psychological distress and quality of life, with the intervention group showing significant improvement compared to the control group. These findings are in line with the evidence showing that psychological distress or mood could be a determinant of the quality of life of patients with cancer^41,42^.

In our study, the intervention participants’ greater improvements in psychological distress and quality of life could be explained through the process indicators (Appendix III). The acceptance and implementation of early palliative care were higher in the intervention group as was evident through the results of process indicators: the percentage of patients who were aware of the information of the cancer journey (Intervention group 74.5% vs control group 63.6%), patients who were following a diet plan (Intervention group 69.1% vs control group 16.4%), patients who received the financial allowance (Intervention group 70.1% vs control group 34.5%), patients who had their pain managed (Intervention group 92.3% vs control group 67.3%), patients who followed any type of mindful-based practice daily (Intervention group 78.2% vs control group 47.2%). The slight improvements seen in the control group’s main outcomes could be also supported by the results of the process indicators since for each component of the novel palliative care intervention package there is some level of reception in the control group as well through the routine healthcare system in Sri Lanka where early palliative care is provided in an ad hoc manner.

Concerning the quality of life dimensions, the results demonstrated a significant improvement in the symptom status in the intervention group. This is an uncommon finding since a systematic review of palliative care effectiveness analyzed 14 studies that measured symptom intensity using a variety of scales, only 1 showed an improvement^39^. It is possible that the current study finding could be due to the participants being just after the diagnosis and the initial cancer treatment itself contributed to the improvements of the symptom status. However, the current study findings can be supported by a randomized control trial conducted on patients with advanced cancers providing early palliative care showed that at 4 months post-intervention, there were significant differences in change scores of quality of life, symptom severity, and satisfaction with care. This study has more power in the findings being a randomized control trial. Although the target group was different in the two studies, since this study focused on patients with advanced cancers of lung, gastrointestinal, genitourinary, breast, and gynaecological the current study was only on patients with oral cancer, the findings are in line^17^.

There are several studies which have failed to show an impact on the quality of life by palliative care interventions. This may be due to a lack of comprehensiveness in the study design, and the presence of only one or two of the domains of the quality of life in the intervention^43-45^. A study carried out on patients with head and neck cancer in India did not show significant improvements in the quality of life after 1 month, 2 months, and three months of randomization^21^. The author attributes this result to the fact that the control group also received a comparable level of palliative care through the routine healthcare system. Also, another reason for the different findings could be that the participants of this study had been patients with stage IV disease or recurrence not amenable to curative treatment and planned for palliative intent chemotherapy whereas the current study included patients presenting at all stages of the disease.

### Clinical Implications

The early palliative care intervention package developed by the present study could be incorporated into the existing health system as a national-level palliative care programme for patients with oral cancer. The newly appointed Public Health Nursing Officers could link the Tertiary Care Hospital and the community level to improve the psychological distress and quality of life of the patients with oral cancer.

### Strengths and limitations of the study

Our study has several strengths to be acknowledged. The response rate being 100% is an outstanding finding of this study and could be attributed to several factors. The participants are just after the diagnosis. Therefore, the enthusiasm for follow-up care is higher than the patients who are in the latter part of the cancer journey. Stringent measures, such as aligning appointments with clinic visits and persistent phone reminders, would have also minimized dropouts. Additionally, the provision of ‘free’ healthcare, lower patient empowerment, and the doctor-patient power dynamic in Sri Lanka likely improved patient compliance.

We acknowledge a few limitations in the study. The selection of a quasi-experimental study has introduced its own biases, however being the first study on early palliative care provision for patients with oral cancer in Sri Lanka this could be considered as a rational starting point for further research in this arena. We used the presence of an accessible Public Health Nursing Officer to allocate the participants to the intervention and control groups. A Public Health Nursing Officer is attached to a Healthy Lifestyle Clinic at a Base Hospital in Sri Lanka, and he/she is responsible for the same area covered by the Base Hospital. Therefore, being away from the hospital affects receiving the services which means there is a bias introduced based on the socio-economic-status of the patient.

The measures used to assess the outcome indicators are both validated to Sri Lanka. However, the process indicators developed in the current study only considered content and face validation. Therefore, this could be a limitation and it is recommended to follow a comprehensive validation for the process indicators in future studies.

We identify the exclusion of any patient who was unable to communicate in Sinhala as a limitation since Sri Lanka is a country with multi-ethnicity the ethnic and racial representation would not have been present. Hence, we recommend the necessity of replicating this study with more diverse populations.

## Conclusions

The novel early palliative care intervention package was effective in improving psychological distress and the quality of life of patients with oral cancer in Sri Lanka. The Public Health Nursing Officers could possibly link the Tertiary Care Hospital and the ground level to improve the quality of life of patients with oral cancer in Sri Lanka.

## Patient and public involvement

During the development of this intervention the inputs of the patients were taken into consideration by conducting in-depth interviews with the patients. The findings of the study will be disseminated through the patients with oral cancer who come to the tertiary cancer care units in Sri Lanka.

## Author contributions

Conception and design: Nadisha Ratnasekera, Irosha Perera, Pushpakumara Kandapola Arachchige

Literature review: Nadisha Ratnasekera, Irosha Perera, Pushpakumara Kandapola Arachchige, Sumeth Perera

Data analysis: Nadisha Ratnasekera, Irosha Perera, Pushpakumara Kandapola Arachchige, Sumeth Perera, Jenny Lau, Camilla Zimmermann

Data interpretation: Nadisha Ratnasekera, Irosha Perera, Pushpakumara Kandapola Arachchige, Sumeth Perera, Jenny Lau, Camilla Zimmermann

Manuscript drafting: Nadisha Ratnasekera

Manuscript revision and approval of final version: Nadisha Ratnasekera, Irosha Perera, Pushpakumara Kandapola Arachchige, Sumeth Perera, Jenny Lau, Camilla Zimmermann

## Acknowledgments

The authors express their gratitude to the consultants in the clinical units for their invaluable assistance in data collection. Special acknowledgment is extended to Dr. Suraj Perera at the National Cancer Control Programme, Sri Lanka, for his support in coordinating the Public Health Nursing Officers. Additionally, we would like to acknowledge Le Lisa, the biostatistician at Princess Margaret Cancer Hospital, for her valuable contribution to the data analysis.

## Data Availability Statement

The data that support the findings of this study are available from the corresponding author upon reasonable request.

